# Radiologic, Pathologic, and Deep Learning Predictors of Response to Immune Checkpoint Blockade in Renal Cell Carcinoma Patients Undergoing Post-Treatment Nephrectomy

**DOI:** 10.1101/2025.11.19.25340277

**Authors:** Payal Kapur, Alana Christie, Vipul Jarmale, Farzad Tahmasebi, Bingqing Xie, Jeffrey Miyata, Dinesh Rakheja, Jeffrey A. Cadeddu, Vitaly Margulis, Hans Hammers, Tian Zhang, Ivan Pedrosa, James Brugarolas, Satwik Rajaram

**Affiliations:** Departments of Pathology; Departments of Urology; Kidney Cancer Program at Simmons Comprehensive Cancer Center; Departments of Radiology; Departments of Bioinformatics; Hematology-Oncology Division of Internal Medicine, University of Texas Southwestern Medical Center, Dallas, TX, 75390

**Keywords:** renal cell carcinoma, predictive biomarkers, prognosis, ccRCC, clear cell renal cell carcinoma, immunotherapy, neoadjuvant, cytoreductive therapy

## Abstract

**Background:** Response assessment of primary kidney tumors in the consolidation cytoreductive and neoadjuvant settings offers a unique opportunity to inform postoperative adaptive treatment strategies. Yet, systematic analyses evaluating radiology, pathology, and machine learning are lacking.

**Methods:** We retrospectively identified consecutive renal cell carcinoma (RCC) patients with locoregionally advanced or metastatic RCC who received at least one cycle of ICI-containing doublet therapy prior to nephrectomy at the UTSW Kidney Cancer Program (2017–2024). Radiologic and pathologic features were centrally reviewed and correlated with clinical outcomes: freedom from start of next systemic therapy (FFNT) in cytoreductive patients and metastasis-free survival (MFS) in neoadjuvant patients. Pathologic response to ICI results in tumor cell death and fibrosis creating hypocellular areas and increased immune infiltrate, features we utilized to build Deep learning (DL) models. We leveraged DL models to validate pathologist-assessed regression objectively and quantitate immune infiltrate.

**Results:** Among 99 patients (cytoreductive nephrectomy {CN}, n=66; neoadjuvant nephrectomy {NaN}, n=33), radiologic tumor shrinkage ≥30% (*p*=0.0036) and the extent of ICI-induced pathologic regression as assessed by central review (HR 0.97; CI 0.95-0.99; *p*=0.0023) and by DL (HR 0.96; CI 0.93-0.99; *p*=0.0041), but not coagulative necrosis, were significantly associated with prolonged FFNT, with similar trends in neoadjuvant cohort. Multivariable Cox regression analyses showed pathologic regression, DL-derived extent of immune infiltrate and tumor largest dimension at nephrectomy to be independent predictors of FFNT.

**Conclusions:** This study provides, for the first time, an integrated, quantitative framework of post-ICI response in RCC. Our data suggests that immune-mediated pathologic regression changes differ from coagulative necrosis, an indicator of poor prognosis. Our findings provide a blueprint for complementary role of radiology and pathology evaluation of post ICI-nephrectomy specimens that if validated prospectively, could guide adaptive approaches and clinical trial design for ICI-based therapies in kidney cancer and beyond.

## Introduction

Management of advanced and metastatic renal cell carcinoma (RCC) has undergone major changes over the past two decades (1). The role of cytoreductive nephrectomy (CN), i.e. surgical resection of primary tumor in the setting of metastatic disease, has been redefined. Clinical trials such as CARMENA and SURTIME demonstrated non-inferiority in overall survival (OS) with systemic therapy alone compared to nephrectomy and systemic therapy (with sunitinib) (HR 0.89; 95% CI 0.71-1.10) (2, 3). Post-hoc analyses suggested that a subset of patients, particularly those with favorable risk profiles and limited metastatic burden, derived benefit from delayed CN, shifting the cytokine era standard of care from upfront nephrectomy to prioritizing systemic therapy and selectively delaying nephrectomy (4).

The introduction of immune checkpoint inhibitors (ICIs) was a landmark advancement in the treatment of advanced RCC. Doublet systemic therapy with inhibitor of programmed cell death protein 1 (PD-1) or its ligand (PD-L1) in combination with either a cytotoxic T-lymphocyte antigen 4 (CTLA-4) inhibitor (ipilimumab) or a vascular endothelial growth factor (VEGF) tyrosine kinase inhibitor (TKI) has significantly prolonged survival, offering durable responses in a subset of metastatic RCC patients, who historically faced dismal prognosis (5–7). More recently, pembrolizumab, a PD-1 inhibitor, was approved in the adjuvant setting (8). Preclinical and phase 2 studies further suggest that neoadjuvant administration of ICI can be superior to adjuvant therapy (9, 10). This may, in part, be attributed to the preservation of tumor-reactive T cells within the intact primary tumor microenvironment. Removal of the primary tumor prior to immune priming may, conversely, deplete this critical reservoir of tumor-specific T cells and limit the breadth of the antitumor response. In RCC, emerging retrospective series suggest that a subset of patients benefit from upfront ICI therapy with deferred CN (11, 12). Building on these findings, there is growing interest in extending ICI-based approaches into the neoadjuvant setting for patients with high-risk localized or locally advanced RCC with the aim of reducing recurrence and prolonging OS.

A major concern with expanding the use of perioperative systemic therapy is the exposure of patients to potential toxicities (including financial) without clear individualized benefit, as many may be cured with surgery alone. This underscores the need to better define which patients derive benefit and which exhibit resistance, thereby informing postoperative adaptive treatment strategies. The NADINA trial in melanoma exemplifies this paradigm: neoadjuvant ipilimumab plus nivolumab (Ipi+Nivo) followed by response-adapted adjuvant therapy significantly improved outcomes compared to adjuvant-alone therapy (13). Importantly, patients who achieved a major pathologic response after just two cycles of neoadjuvant therapy could be de-escalated, omitting adjuvant therapy without compromising survival.

One of the main barriers to integrating such adaptive neoadjuvant ICI therapy into RCC care is the lack of reliable biomarkers and early surrogates of response. Radiologic evaluation, while noninvasive, has limitations. Comparison of sequential scans can accurately measure tumor shrinkage but cannot reliably differentiate viable from non-viable tumor, particularly at the microscopic level. Further, radiologic staging remains suboptimal for detecting T3a disease (e.g., renal sinus or early renal vein invasion) resulting in over staging cT2 tumors (14). In contrast, though pathology cannot assess longitudinal shrinkage, pathologic assessment of treated tumors provides cellular level insights into tumor viability and response, including extent of immune infiltrate. In addition, it also provides adequate tissue for biomarker development. In other cancer types, pathologic assessment has successfully served as an early surrogate to predict disease-free survival and OS (15, 16). However, no standardized guidelines currently exist for evaluating nephrectomy tumor samples post ICI, and the practical challenges of sampling these large tumors (unlike other cancer types) make whole tumor/slab submission unrealistic in routine practice.

Workflows for evaluating response in histology samples in clinical trials vary, typically involving evaluation at local sites, with central review to establish consensus. Central reviews generally allow for a consistent analysis and minimize inter reader variability. Today, automated machine learning (ML)-based approaches can enable scalable standardized analyses of pathologic features such as quantify extent of nonviable tumor (pathologic response), presence of coagulative tumor necrosis, and inflammation, complementing visual methods for more accurate and consistent pathology response assessment of neoadjuvant specimens (17). To date, this has not been explored in RCC.

In this study, we sought to address these challenges by identifying surrogate biomarkers for early response to pre-nephrectomy frontline ICI doublet therapies. To minimize interobserver variability and enable scalability, we leverage digital pathology-based deep-learning models. We evaluated the association of both visual and digital response assessments with clinical outcomes. Furthermore, we explored whether biomarkers identified in the cytoreductive setting could be extrapolated to the neoadjuvant context, where immune dynamics may differ due to the absence of distant metastases and shorter therapy duration. This work aims to define practical, deliverable endpoints that can inform future clinical trials and ultimately guide clinical practice.

## Material and Methods

### Patient selection criteria

This retrospective study was conducted with approval by the Institutional Review Board (STU 022015-015). Eligible patients included those diagnosed with metastatic or locally advanced RCC treated at UT Southwestern (Dallas, Texas), who received at least one cycle of ICI-based frontline doublet therapy prior to undergoing partial and/or radical nephrectomy. Acceptable ICI regimens included PD-1 or PD-L1 therapies, administered in combination with CTLA-4 blockage or VEGF-TKIs. Patients who received multiple lines of systemic therapy before nephrectomy or who underwent nephrectomies at outside institutions were excluded.

### Data collection

Patient identification and data abstraction were performed using our institutional kidney cancer database (Kidney Cancer Explorer {KCE}), a HIPAA-compliant data management platform. KCE integrates data from the electronic medical record (EPIC) through automated pipelines, including custom natural language processing (NLP) and more recently large language models (LLMs), for extracting pathology data from free-text reports (18, 19). Structured variables such as type of systemic therapy are captured using SQL and python scripts. Documentation of metastases is based on histology confirmation when available, or otherwise on the initiation of systemic therapy or radiation. Data quality controls include automated discrepancy flags and manual review. Non-discrete clinical fields were supplemented through manual chart review.

### Radiologic analysis

Patients are routinely followed up after diagnosis with physical examination, laboratory studies, and chest computerized tomography (CT), and abdominal/pelvic CT or magnetic resonance imaging (MRI) every 6-16 weeks for metastatic disease, and every 3-12 months for non-metastatic disease. Standard imaging obtained immediately before initiation of ICI therapy and prior to nephrectomy were reviewed (F.T., 7 years of experience) under supervision (I.P., 25 years of experience). Three orthogonal dimensions of the primary tumor were recorded in using axial, coronal, and sagittal images. Oblique measures to capture the longest dimension were allowed. Tumor shrinkage was calculated by comparing largest dimension measurements prior to initiation of therapy and pre-nephrectomy timepoints.

### Pathologic analysis

Hematoxylin and eosin (H&E) stained clinical slides and gross images of the nephrectomy specimens were retrieved. Currently no standardized guidelines exist for evaluating and reporting post-ICI nephrectomy findings. At our institution, the tumor bed is photographed, and representative sections are obtained from all macroscopically different areas. Sampling is based on tumor size- at least 1 section per centimeter of tumor dimension, with inclusion of all heterogeneous areas. If no viable tumor is identified, the entire tumor bed is submitted for microscopic evaluation. Representative sections of thrombus and metastases when present are also similarly submitted. Tumors less than 3 cm are entirely submitted for evaluation.

Pathology reports follow the College of American Pathologists (CAP) cancer template, including documentation of coagulative tumor necrosis (i.e., presence must be documented, but extent is optional). All clinical H&E slides were available for 92 nephrectomies and were subjected to central microscopic review (P.K., 25 years of experience). For 7 cases, without available slides, the extent of viable tumor was inferred from the pathology report.

### Digital Pathology

<u>Acquisition</u>: For each patient, two representative H&E slides were scanned at 40x magnification using a Leica GT450 DX scanner. The most representative tumor sections, as determined during routine pathologic evaluation, were included. If only one slide has originally been selected, an additional slide was identified during central review to ensure that the sampled slides collectively captured the extent of viable tumor. All subsequent analyses were performed on these images at 20x magnification.

<u>Region Identification</u>: Two representative digitalized whole slide images (WSI) were available for digital pathology evaluation from 94 cases. An in-house region classification model was used to classify 224 pixels (∼112 micron) H&E image patches at 20x into the following classes: background, blood, tumor, normal kidney, stroma, fat, immune-rich areas, areas of coagulative necrosis, and other (typically hyalinization) areas. This model was applied across each WSI in a grid with 64 pixels spacing to produce a low-resolution regional mask. The classification for each point on this grid was determined by analyzing its surrounding 224 pixels tissue patch. These regional classifications were then refined in two ways. First, the GrandQC (20) model was applied to identify and exclude staining and tissue artifacts by reclassifying them as background. Second, a pathologist (PK) blinded to clinical data performed a manual review to exclude stromal regions outside the tumor region via manual annotation. After exclusion of the background areas, fat and blood regions, the final outputs were masks defining the total tumor-associated tissue and its subregions of coagulative necrosis.

<u>Nucleus classification</u>: We utilized a previously developed U-Net-based semantic segmentation model to classify pixels in 20x H&E images (21, 22). This model was trained using ground truth from matched IHC stained WSI to identify pixels as background or belonging to immune (CD8+), tumor (PAX8+), endothelial (ERG+), or other cells. To convert these pixel-level calls to nucleus-level counts, we identified boundaries of individual nuclei using the public HoverNext model (23). Each nucleus instance found by HoverNext was then assigned to one of the aforementioned classes based on a majority vote of its constituent pixels. For downstream analysis, we restricted analysis to the previously defined tumor-associated tissue and used the resulting nuclear areas as well as the counts of total and immune nuclei.

<u>Hypocellular-area identification:</u> A feature of ICI response is hypocellular areas either in the form of cell death or fibrosis. We identified these hypocellular areas based on local nuclear density. To quantify this, we tessellated the WSI using 224 pixels patches with a 64 pixels stride (consistent with the regional analysis) and classified any patch with less than 3.75% nuclear pixel area as "hypocellular". This cutoff was chosen based on manual review of a set of WSI with no viable tumor using a range of low-density cutoffs to best select the tumor response area.

<u>Response metrics:</u> From these analyses, we derived three quantitative histologic features: (1) Tumor bed region: calculated as the total tumor-associated tissue area per WSI after removal of fat, benign kidney and perinephric stromal regions. (2) The proportion of coagulative necrosis: calculated as the area of coagulative necrosis divided by the tumor bed area. (3) The proportion of responsive tissue: calculated as the hypocellular area minus the coagulative necrosis area, divided by the tumor bed area. (3) The immune cell percentage: calculated as the proportion of all nuclei classified as immune cells. To obtain patient-level scores, the numerators and denominators for each metric were separately aggregated across all of a patient’s slides before the final ratio was calculated.

### Clinical endpoints

Clinicopathologic and follow-up data were extracted from KCE last updated on August 11th, 2025. Primary outcomes included metastasis and death. For the cytoreductive nephrectomy (CN) cohort the primary outcome was time from nephrectomy to start of next systemic therapy (freedom from start of next therapy- FFNT), or censored at last follow-up/ death, whichever came first. For the neoadjuvant cohort, the endpoint was metastasis-free survival (MFS) defined from the date of nephrectomy to the date of first documented metastasis, or censored at last follow-up/ death, whichever came first.

### Statistical methods

All features were tabulated and summarized with event frequencies, percentages, or median and interquartile ranges (IQR), as appropriate. Survival analyses for FFNT and MFS were estimated by Kaplan-Meier methods, and Cox proportional hazards models were applied to assess association between features and outcomes, including in multivariable models. Hazard ratios (HR) with 95% confidence interval (CI) and *p*-value were reported. Unless otherwise specified, *p*-values are two-sided and unadjusted for multiple testing. All statistical analyses were completed using SAS 9.4 (SAS Institute Inc., Cary NC).

## Results

### Baseline characteristics of Cytoreductive Nephrectomy (CN) and Neoadjuvant Therapy prior to Nephrectomy (NaN) cohorts

A total of 99 patients with advanced or metastatic RCC who had ICI-based doublet therapy as the first systemic therapy and received at least 1 cycle, were included in this study. The cohort comprised 66 metastatic RCC patients who underwent CN and 33 patients with non-metastatic, locally advanced RCC who underwent neoadjuvant therapy prior to nephrectomy (NaN). Although neoadjuvant therapy is not standard of care for RCC, the documented rationale for using systemic therapy prior to nephrectomy, as noted in the clinical records included: (1) downstaging an unresectable tumor to enable resection; and (2) facilitating partial nephrectomy in the setting of renal insufficiency or multiple/bilateral tumors. The median age at start of systemic therapy (62 years), baseline demographics, as well as median follow-up were comparable (2.5 [1.0-3.6] years in CN vs. 2.1 [1.2-4.1] years in NaN) (**Suppl. Table 1)**. Key systemic therapy related differences between CN and NaN cohorts included: (1) shorter duration of pre-nephrectomy ICI therapy in the NaN cohort (6.1 [3.7-9.5] month in CN vs. 4.9 [3.4-6.2] months in NaN); (2) a higher proportion of Ipi+Nivo use in CN patients [59% vs. 24%] (**Suppl. Table 1)**. The observed differences in treatment duration and regimen composition between the CN and NaN cohorts highlight the importance of accounting for therapeutic context when evaluating biomarkers of early response.

### Radiologic reduction in primary tumor size with ICI therapy correlates with clinical outcome in the CN and NaN cohorts

To assess temporal change in tumor size prior to nephrectomy, we turned to radiologic studies. Baseline radiologic primary tumor dimensions were similar across cohorts (median 10.4 cm [8.0-12.4] in CN vs. 8.8 cm [7.1-11.8] in NaN), as was the distribution of clinical T substage (cT3/4: 62% in CN vs. 73% in NaN) **(Suppl. Table 1)**. When evaluating associations with outcomes, neither baseline clinical T substage (cT) nor the IMDC risk group reached statistical significance for FFNT or MFS in either cohort (**Table 1 and 2**). However, the hazard ratios suggested a directional but non-significant trend towards poorer outcomes with higher T stage or IMDC risk, raising the possibility of a modest effect not fully captured due to sample size limitations. This underscores that in the context of ICI therapy, traditional anatomic and clinical risk parameters may be less predictive, and we turned to assess dynamic treatment-associated changes.

**Table 1.** Cox proportional hazards analysis of time to starting new systemic therapy after nephrectomy, in patients receiving cytoreductive nephrectomy after IO

| N=66 | N (%) or<br>Median (IQR) | N<br>Events | Hazard Ratio<br>(95% CI) | P |
| --- | --- | --- | --- | --- |
| Age at Systemic Treatment | 62.5 (57-67) | 25 | 1.01 (0.96, 1.05) | 0.8 |
| Sex |  |  |  |  |
| Female | 16 (24%) | 1 | Reference | 0.055 |
| Male | 50 (76%) | 24 | 7.12 (0.96, 52.77) |  |
| Race |  |  |  |  |
| White | 55 (83%) | 19 | Reference | 0.11 |
| Non-White | 11 (17%) | 6 | 2.17 (0.85, 5.55) |  |
| Ethnicity |  |  |  |  |
| Non-Hispanic | 53 (84%) | 22 | Reference | 0.3 |
| Hispanic | 10 (16%) | 2 | 0.46 (0.11, 1.98) |  |
| Oncology |  |  |  |  |
| Clinical T stage at Dx |  |  |  |  |
| 1 | 10 (16%) | 5 | Reference | 0.6 |
| 2 | 13 (21%) | 3 | 0.59 (0.14, 2.48) |  |
| 3 | 30 (49%) | 14 | 1.36 (0.49, 3.79) |  |
| 4 | 8 (13%) | 2 | 1.09 (0.20, 5.80) |  |
| IMDC risk group |  |  |  |  |
| Intermediate risk | 28 (74%) | 9 | Reference | 0.4 |
| Poor risk | 10 (26%) | 5 | 1.62 (0.54, 4.84) |  |
| Type of ICI |  |  |  |  |
| ICI+ICI | 39 (59%) | 15 | Reference | 0.7 |
| ICI+TKI | 27 (41%) | 10 | 1.20 (0.53, 2.71) |  |
| Months from start of ICI to Nx | 6.1 (3.7-9.5) | 25 | 0.93 (0.86, 1.00) | 0.064 |
| Continued ICI post-Nx |  |  |  |  |
| Yes | 25 (54%) | 8 | Reference | 0.9 |
| No | 21 (46%) | 10 | 1.04 (0.41, 2.64) |  |
| Months on ICI post-Nx (n=46) | 1.4 (0.0-9.6) | 18 | 0.94 (0.88, 1.01) | 0.081 |
| Radiology |  |  |  |  |
| Size prior to ICI (cm) (n=49) | 10.4 (8.0-12.4) | 20 | 1.04 (0.92, 1.17) | 0.6 |
| Size prior to Nx (cm) (n=56) | 8.7 (7.1-11.5) | 22 | 1.13 (0.999, 1.29) | 0.051 |
| Change in size (cm) (n=44) | -1.5 (-3.5-0.0) | 18 | 1.42 (1.11, 1.81) | 0.0051 |
| Change in size (%) (n=44) | -18.7 (-25.8-0.2) | 18 | 1.03 (1.01, 1.06) | 0.0035 |
| Pathology at Nx |  |  |  |  |
| Histological Subtype |  |  |  |  |
| ccRCC | 59 (89%) | 21 | Reference | 0.3 |
| non-ccRCC | 7 (11%) | 4 | 1.78 (0.61, 5.20) |  |
| Sarcomatoid |  |  |  |  |
| Not identified | 49 (79%) | 19 | Reference | 0.4 |
| Present | 13 (21%) | 6 | 1.52 (0.60, 3.83) |  |
| Rhabdoid |  |  |  |  |
| Not identified | 50 (82%) | 18 | Reference | 0.18 |
| Present | 11 (18%) | 7 | 1.82 (0.76, 4.37) |  |
| Coagulative necrosis |  |  |  |  |
| Not identified | 20 (31%) | 7 | Reference | 0.5 |
| Present | 44 (69%) | 18 | 1.33 (0.55, 3.19) |  |
| Extent of necrosis (%) (n=53) | 10 (0-40) | 23 | 0.99 (0.98, 1.01) | 0.3 |
| Grade |  |  |  |  |
| G2 | 6 (10%) | 0 | * | 0.8 |
| G3 | 28 (48%) | 12 | Reference |  |
| G4 | 24 (41%) | 12 | 1.35 (0.60, 3.01) |  |
| Pathological T stage at Nx |  |  |  |  |
| ypT0 | 6 (9.1%) | 0 | * | 0.3 |
| ypT1 | 7 (11%) | 1 | Reference |  |
| ypT2 | 5 (7.6%) | 1 | 1.40 (0.09, 22.48) |  |
| ypT3 | 40 (61%) | 18 | 4.50 (0.60, 33.81) |  |
| ypT4 | 8 (12%) | 5 | 7.55 (0.87, 65.32) |  |
| Change in T stage |  |  |  |  |
| Decreased | 16 (28%) | 2 | Reference | 0.15 |
| Unchanged | 28 (49%) | 12 | 4.01 (0.90, 17.94) |  |
| Increased | 13 (23%) | 8 | 4.51 (0.95, 21.36) |  |
| Pathological N stage at Nx |  |  |  |  |
| ypN0/X | 56 (85%) | 19 | Reference |  |
| ypN1 | 10 (15%) | 6 | 2.64 (1.05, 6.64) | 0.039 |
| Largest dimension (cm**) | 8.1 (6.5-10.7) | 25 | 1.17 (1.07, 1.28) | 0.0010 |
| Volume (cm<sup>3</sup>*) | 289 (158-662) | 25 | 1.004 (1.001, 1.01)*** | 0.018 |
| **Pathology Central Review** |  |  |  |  |
| Coagulative necrosis |  |  |  |  |
| Not identified | 23 (38%) | 6 | Reference | 0.068 |
| Present | 37 (62%) | 18 | 2.37 (0.94, 5.99) |  |
| Pathologic regression (%) (n=60) | 30 (15-70) | 24 | 0.97 (0.95, 0.99) | 0.0023 |
| Viable tumor (%) | 60 (20-80) | 25 | 1.02 (1.01, 1.04) | 0.0025 |
| Viable tumor volume (cm<sup>3</sup>) | 137 (38-324) | 25 | 1.01 (1.003, 1.01)*** | 0.0013 |
| **Digital Pathology (n=64)** |  |  |  |  |
| Immune cells (%) | 31.7 (21.5-39.5) | 23 | 0.97 (0.93, 1.003) | 0.069 |
| Coagulative necrosis (%) | 0.2 (0.0-2.8) | 23 | 0.98 (0.91, 1.05) | 0.5 |
| Pathologic regression (%) | 29.1 (15.8-43.2) | 23 | 0.96 (0.93, 0.99) | 0.0041 |

\* No events occurred in this level.
\*\* Includes necrotic/regression dimensions, not just viable tumor dimensions. Largest lesion taken for multifocal tumors.
\*\*\* Hazard ratio per increase of 8 cm<sup>3</sup>.
ccRCC: clear cell renal cell carcinoma; Dx: diagnosis; ICI: immune checkpoint inhibitor; IMDC: International Metastatic Renal Cell Carcinoma Database Consortium; Nx: nephrectomy; TKI: tyrosine kinase inhibitor. Numbers are specified in parenthesis when data is missing.

Paired radiologic images at both pre-ICI therapy and pre-nephrectomy time points were available for 73.3% (44 in CN and 29 in NaN respectively), enabling accurate longitudinal comparison (**Table 1 and 2**). Both cohorts demonstrated measurable primary tumor shrinkage prior to nephrectomy (CN: -1.5 cm [-3.5 - -0.0]; NaN: -1.3 cm [-2.3 - -0.6]) (**Suppl. Table 1, Suppl. Figure 1**). Tumor shrinkage was increased for ICI+TKI as compared to ICI+ICI (-1.6 cm vs. -1.1 cm) when both cohorts were evaluated together, but not significantly different (*p*=0.9). In both the CN and NaN cohorts, absolute reduction and percentage change in radiologic tumor size significantly correlated with improved FFNT and MFS respectively (all *p*<0.05) (**Table 1 and 2**). Using the RECIST v1.1 criteria of partial response, we found that patients that experienced ≥30% shrinkage in the radiologic longest tumor diameter did not develop progression or metastasis (20.5% in CN and 17.2% NaN) at the time of last follow-up (**Figure 1**). Overall, these results highlight that radiologic tumor shrinkage is a meaningful early indicator of clinical outcome in patients receiving ICI containing doublet therapy.

**Figure 1.**
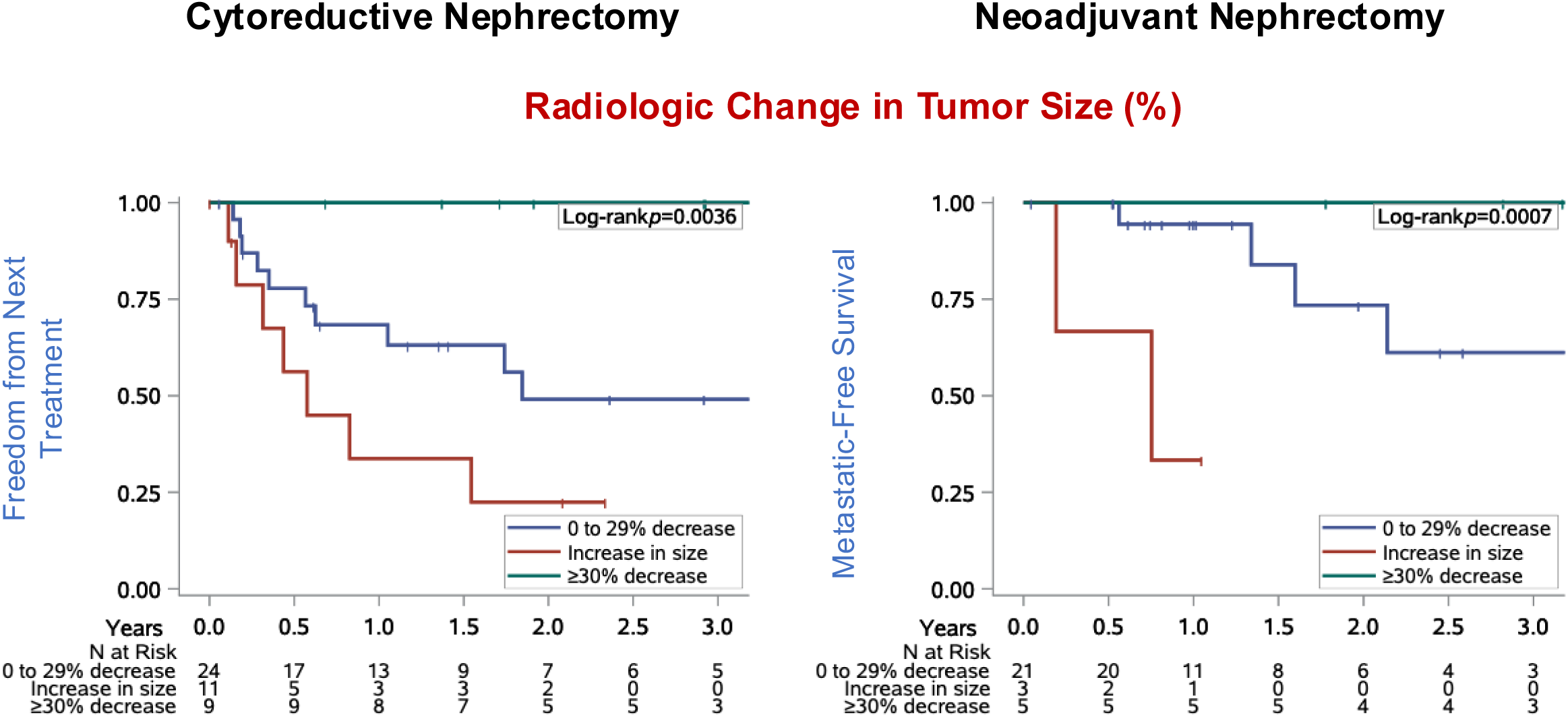
Radiologic evaluation of change in primary tumor dimensions before ICI therapy and prior to nephrectomy in cytoreductive (CN) and neoadjuvant (NaN) cohorts. Kaplan–Meier curves demonstrating that percentage change in radiologic largest tumor dimension significantly correlated with improved FFNT and MFS in the CN and NaN cohorts respectively. *P* values are from log-rank test comparing the three prespecified size change categories as nominal groups; thresholds mirror RECIST (≥30% reduction in size).

### Traditional pathologic features are poor predictors of clinical outcomes

Radiologic and pathologic size estimates can differ slightly, but more importantly renal vein branch involvement, and hilar and perinephric fat infiltration (early pT3a) are often undetected radiologically, leading to radiologic overestimation of pT2 (14). Notwithstanding these limitations, downstaging (defined as a decrease in ypT relative to baseline cT) was observed in 28% of CN and 42% of NaN patients (**Table 1 & 2**). Pathologic features of nephrectomy samples showed that, compared with CN, the NaN cohort included a significantly higher proportion of lower- grade tumors at nephrectomy (**Suppl. Table 1)**. These differences could partly arise from baseline selection differences, as neoadjuvant therapy patients harbor less advanced tumors. Cox proportional hazards analyses of FFNT and MFS revealed trends towards association with pathologic T substage (ypT), histologic subtype, and grade at nephrectomy, although these did not reach statistical significance in either the CN or the NaN cohorts (**Table 1 and 2**). The presence of sarcomatoid features in the residual tumor did not reach significance in the CN cohort but was associated with markedly worse MFS in NaN cohort (**Tables 1 and 2**). In the CN cohort, presence of viable nodal (ypN1) metastasis, largest tumor dimension (as documented in pathology reports), and tumor volume at nephrectomy strongly correlated with shorter FFNT (all p<0.05) (**Table 1**). In the NaN cohort, tumor volume, but not the largest dimension showed a trend toward correlation with MFS, but it did not reach significance, likely due to smaller cohort size (**Table 2**). Overall adverse pathologic features known to predict poor outcomes in treatment-naïve patients showed diminished predictive ability in post ICI treated samples. These observations underscore the need for novel biomarkers, beyond conventional histopathologic variables, to improve prognostication following ICI therapy.

**Table 2.**
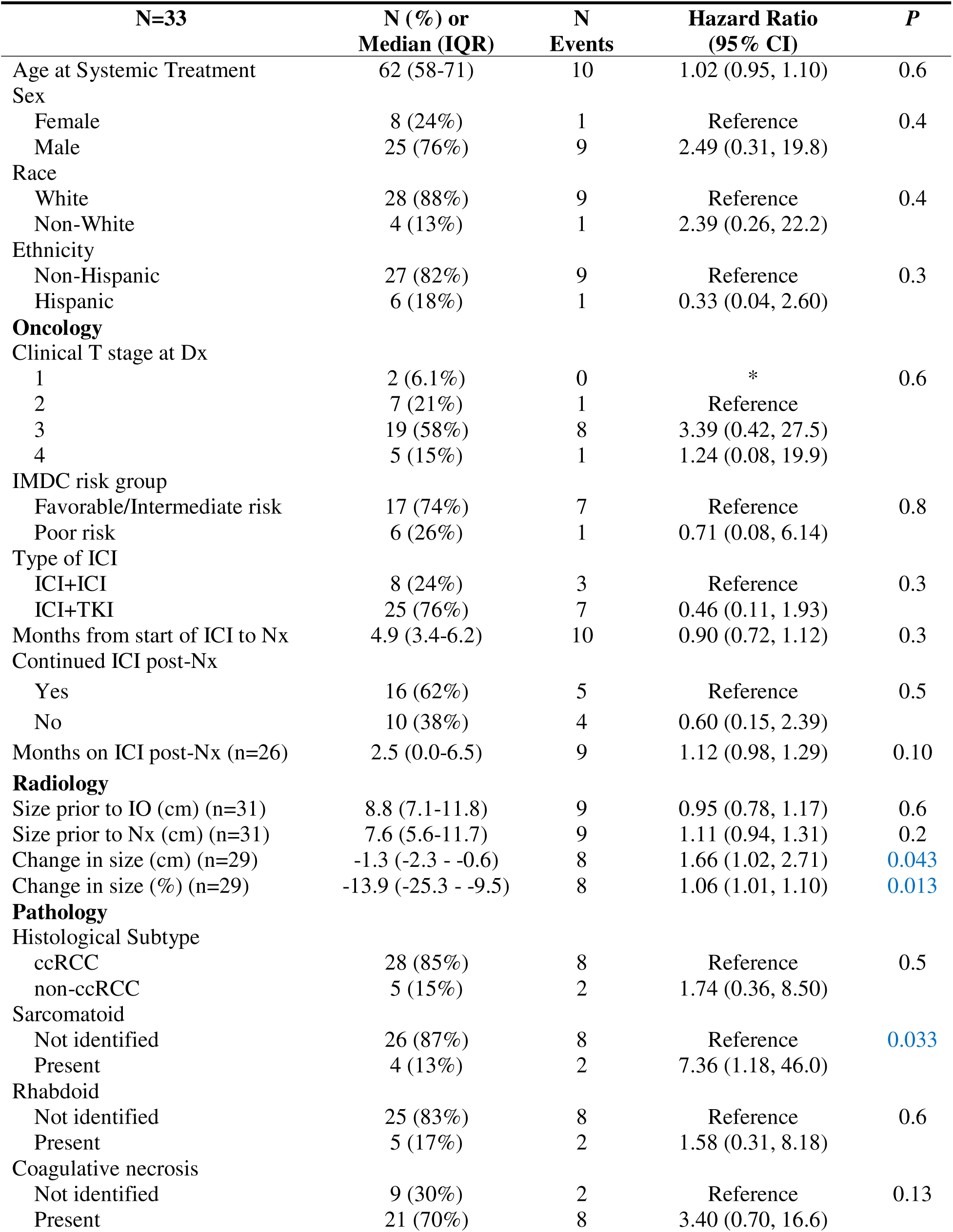
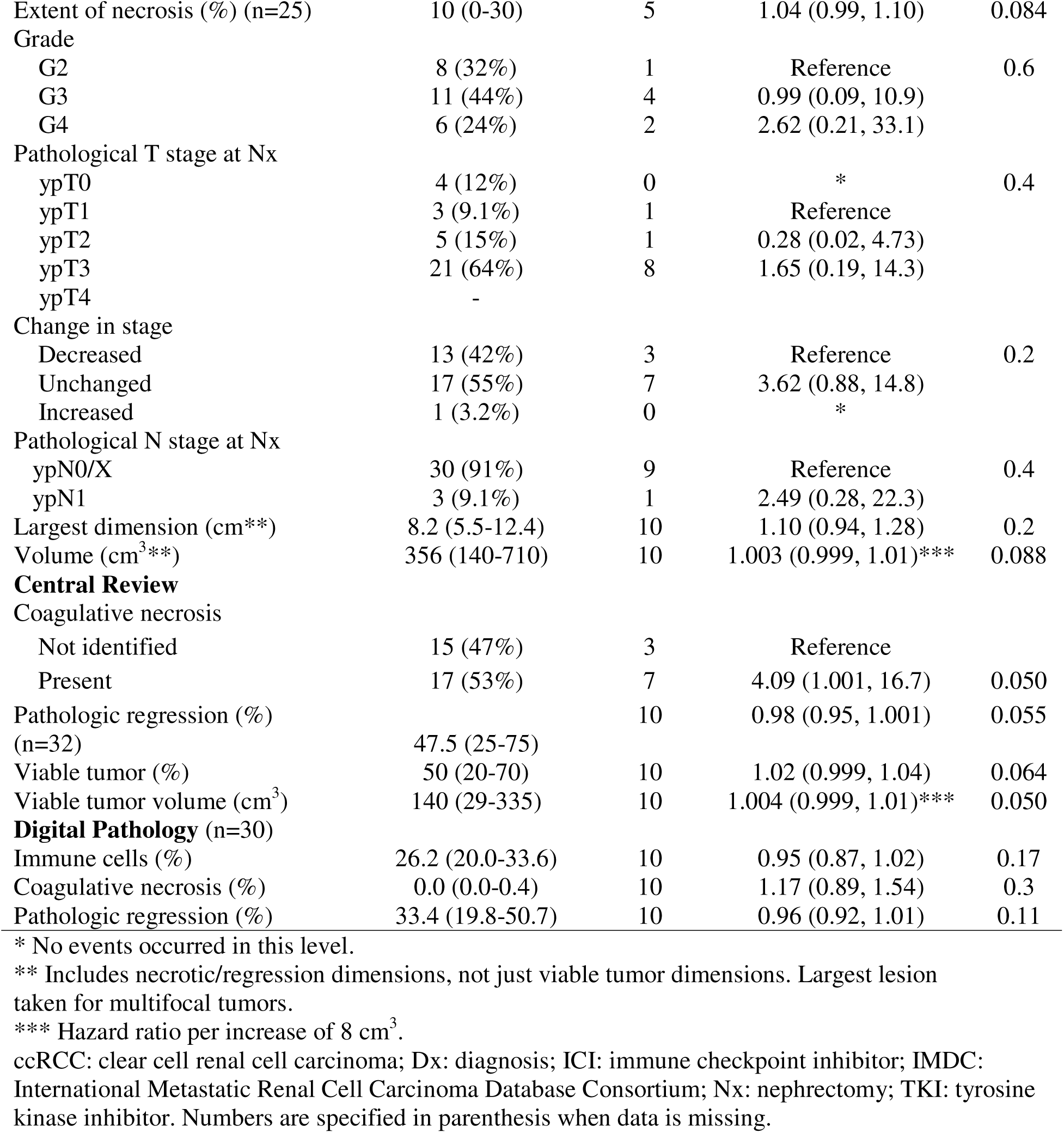
Cox proportional hazards analysis of time to developing metastasis after nephrectomy for patients receiving nephrectomy after neoadjuvant ICI

### Coagulative tumor necrosis remains prognostic post ICI therapy

The current CAP cancer template for nephrectomy specimens requires reporting of presence of tumor necrosis. Notably, necrosis reported in routine pathology records did not correlate with outcomes, reflecting variability in usage; where some pathologists applied strict coagulative necrosis criteria while others incorporated pathologic response such as hyalinization within their estimation of “necrosis” (**Table 1 and 2**). Given these inconsistencies, we centrally reviewed coagulative necrosis and pathologic response metrics separately to capture therapy-related effects and their associations with outcome more accurately. Coagulative tumor type necrosis was defined as geographically well-demarcated, homogenous areas of amorphous tumor cellular debris with preserved cellular outlines and karyorrhectic debris (**Suppl. Figure 2A**). We first assessed the variability in the assessment of coagulative necrosis in the pathology reports compared to central review and the deep learning model that was based on 2 representative WSI. Using Pearson correlation, we found a moderate correlation in the extent of coagulative necrosis across the 3 approaches (r=0.48-0.50) (**Suppl. Figure 2B)**. Similar to untreated patients, presence of coagulative necrosis as assessed through central review showed shorter FFNT and MFS (CN: HR 2.37; 95%CI 0.94-5.99; *p*=0.06 and NaN: HR 4.09; 95%CI 1.001-16.7; 0.05) and this effect was stronger when using log-rank test (*p*=0.060 and 0.037 respectively) (**Suppl. Figure 2C**). Further, the extent of coagulative necrosis significantly correlated with poor prognostic variables such as tumor grade (*p*<0.001) and ypT substage (*p*=0.0031) in the post treatment nephrectomy specimen (**Suppl. Figure 2D)**. Together, these findings suggest that similar to treatment naïve patients, the presence of coagulative necrosis continues to be a poor prognostic feature.

### Pathologic extent of viable tumor and tumor size/volume correlate with outcomes

Next, we centrally quantitated pathologic response metrics which integrate both viable tumor and treatment-associated stromal changes. All forms of non-viable tumor other than coagulative necrosis and acute hemorrhage, were categorized as ICI therapy-induced pathologic regression (iPR) changes encompassing intra- and peri-tumoral fibrosis, hyalinization and ischemic-type necrosis, as described previously and in the methods (24) (**Suppl. Figure 3**). iPR was systematically assessed by reviewing all H&E slides together with corresponding gross images (**Suppl. Figure 3**), blinded to the clinical data. Microscopically, iPR was mapped on each slide and averaged across the entire tumor. Similar to clinical practice, gross photographs were reviewed in parallel to verify that the sampled H&E sections were representative of the overall tumor bed. The estimated range of iPR was adjusted with the narrow bounds observed across slides (<10% variation) when it was felt that areas of regression may not be fully captured histologically.

Quantitative central review estimates of percentage iPR (iPR%) significantly correlated with FFNT in CN patients (HR 0.97; 95% CI 0.95-0.99; *p*=0.0023) and a similar correlation with MFS was seen in the NaN cohort, though did not reach significance (HR 0.98; 95% CI 0.95-1.001; *p*=0.055) (**Tables 1 and 2**). Consistently, Kaplan Meier analyses using the median as threshold demonstrated a statistically significant difference (CN: log-rank p=0.0023; NaN: log-rank p=0.018; **Figure 2A**). All patients with an iPR of >80% are free from events in both cohorts (CN: n = 8; NaN: n = 7). Similar results were seen with viable tumor (VT%) and viable tumor volume (**Tables 1 and 2**). Prior studies have empirically defined major responders and responders as having ≤20% and ≤50% pathologic viable tumor respectively (12). Twenty-seven percent of patients (27/99) achieved VT% of ≤20%. Using this cutoff, Kaplan Meier analyses continued to demonstrate statistically significant difference in FFNT (CN: log-rank p=0.0064; NaN: log-rank p=0.061; **Figure 2B**).

**Figure 2.**
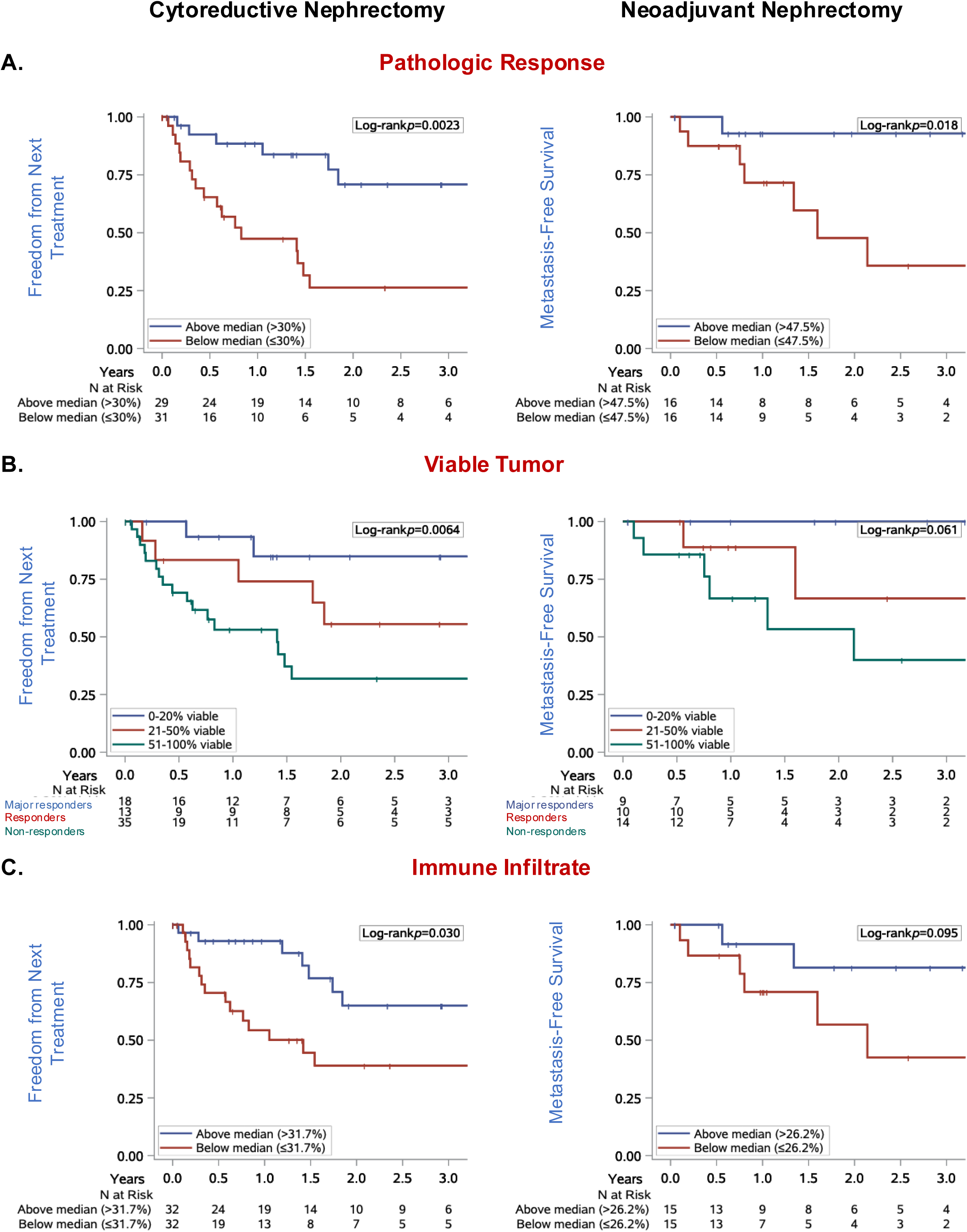
Quantitative pathology and deep learning (DL) correlates of treatment response. (A) Kaplan–Meier curves with stratification based on central review quantification of iPR% using median as cutoffs, showing significant differences in FFNT and MFS. (B) Kaplan–Meier curves stratified by residual viable tumor (VT% ≤20% = major responders; >50% = nonresponders), showing significant differences in FFNT with similar trends for MFS. (B) DL-based quantification of percentage immune infiltrate based on median cutoff for FFNT in CN patients and MFS in NaN patients.

### Deep Learning Model outputs of pathologic regression and extent of immune cell infiltrate correlate with outcomes

We looked for ways to validate the findings from the central review and turned to our DL-model readout of iPR%. Reassuringly, central review and DL model assessments of iPR%, were highly correlated, despite being assessed on only two representative tumor-containing slides (see methods) by the DL model (Pearson correlation coefficient=0.75 (*p*<0.0001); **Suppl. Figure 4A**). By extension, these findings suggest that partial sampling may be sufficient for assessing the extent of viable tumor.

We performed Kaplan-Meier and Cox-proportional hazards analyses. As shown in **Suppl. Figure 4B**, based on two class stratification into the threshold above and below the median (iPR%: 29.1) log-rank analyses demonstrated a statistically significant difference for the CN cohort (*p*=0.0072), but not for the NaN cohort. Of note, the stratification by median cutoff of around 30% by DL is comparable to that observed by central review and in prior studies (25).

Compared to treatment-naïve nephrectomies, we observed a heightened immune infiltrate. We asked if the extent of immune infiltrate in the tumor bed could predict response. Indeed, the percentage of immune infiltrate within the tumor bed, as assessed by the DL model from just two representative slides, strongly correlated with FFNT. Kaplan Meier analyses using the median as threshold demonstrated a statistically significant difference for CN but did not reach significance in NaN (CN: log-rank p=0.03; NaN: log-rank p=0.095). (**Figure 2C**). These results suggest that quantitative assessment of pathologic response and immune infiltrate as assessed objectively from just 2 WSI using DL models provides strong correlation with outcomes following ICI therapy, at least in the CN setting.

### Complete pathologic response predicts excellent prognosis in both CN and NaN

Ten patients achieved a complete pathologic response (absence of any viable tumor) at nephrectomy (6 CN and 4 NaN). Median age was 60.5 years (range, 48–65), with 40% female and 90% White, including 20% Hispanic (**Table 3**). At diagnosis, 60% of patients had cT3/4 tumors, 40% were cN1, and 60% had synchronous metastasis. Patients had intermediate (67%) or poor (33%) IMDC risk disease at the start of therapy. Pretreatment biopsies, available in 7/10 patients, frequently demonstrated high-grade features (57% grade 4, 14% grade 3, 29% grade 2), with sarcomatoid change in 40% and rhabdoid features in 14%. 90% were confirmed to be of ccRCC histology at nephrectomy. Six patients (60%) received IO+TKI and four (40%) received IO+IO. The median time from initiation of immunotherapy to nephrectomy was 8.7 months (range, 3.6–11.5). Radiologically, tumors decreased in size from a median of 9.8 cm before immunotherapy to 6.4 cm before nephrectomy, corresponding to a median reduction of 43.7% in longest dimension. For patients with radiologic images available at both timepoints, 57% (4/7) had ≥30% reduction. At nephrectomy, tumors were predominantly right sided (90%), with a median largest dimension of 6.7 cm and median volume of 162 cm³.

**Table 3.**
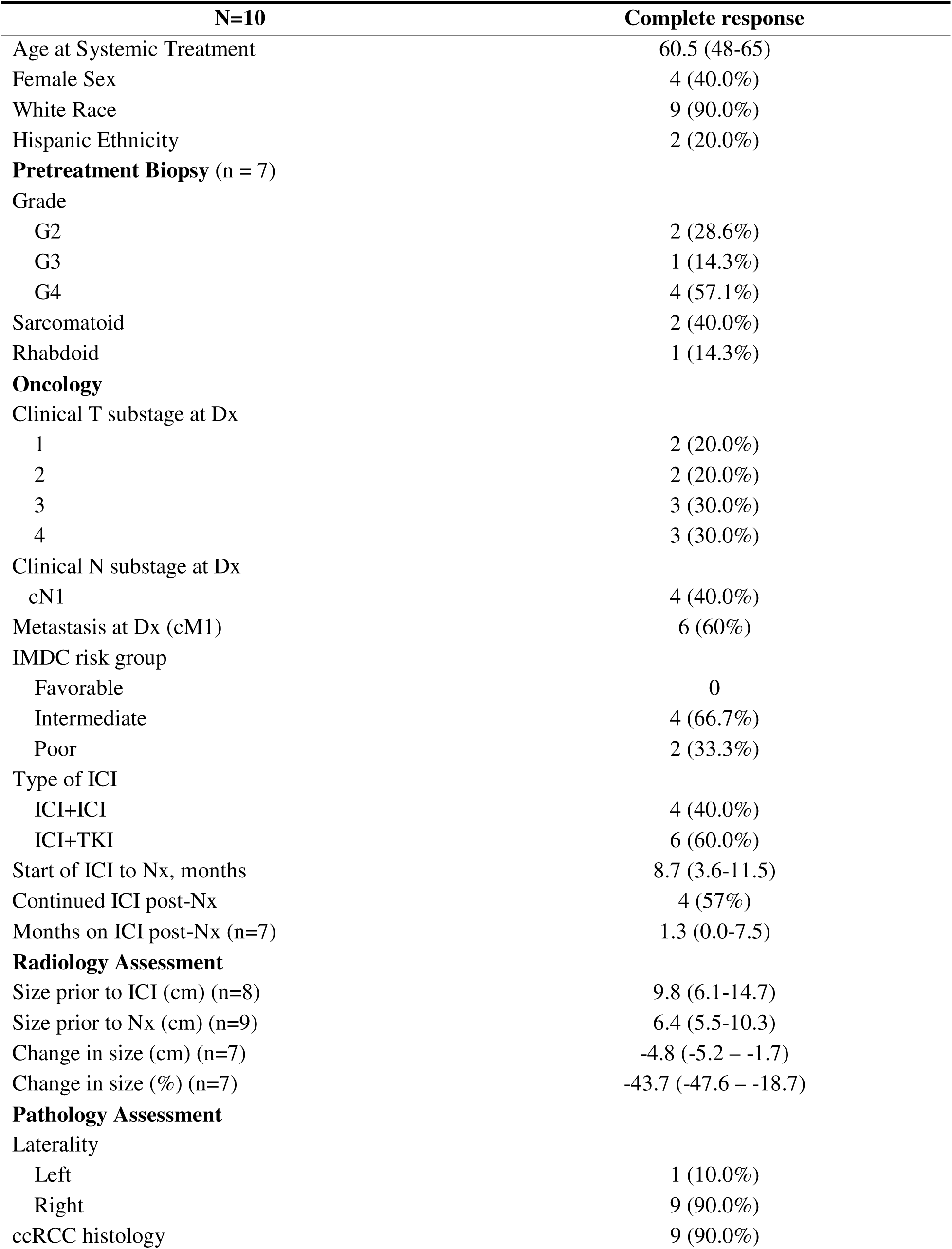
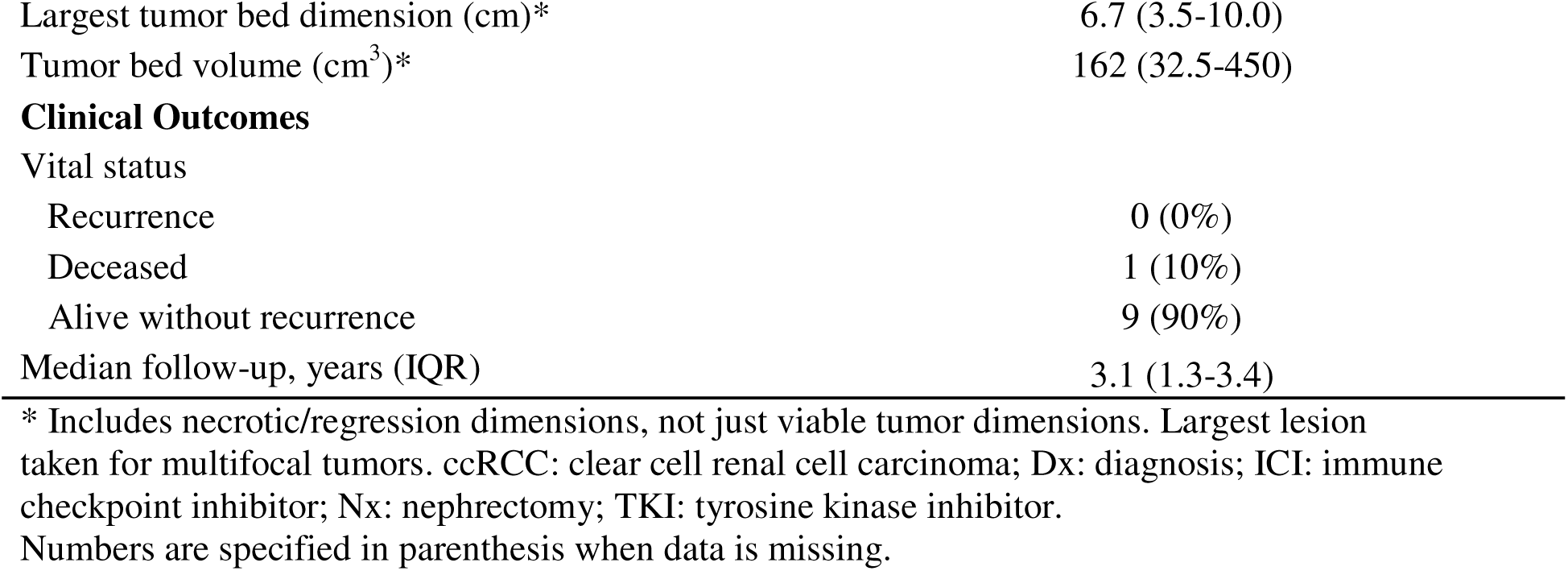
Clinical characteristics of patients receiving cytoreductive or neoadjuvant nephrectomy with complete pathologic response at nephrectomy.

| N=10 | Complete response |
| --- | --- |
| Age at Systemic Treatment | 60.5 (48-65) |
| Female Sex | 4 (40.0%) |
| White Race | 9 (90.0%) |
| Hispanic Ethnicity | 2 (20.0%) |
| **Pretreatment Biopsy (n = 7)** |  |
| Grade |  |
| G2 | 2 (28.6%) |
| G3 | 1 (14.3%) |
| G4 | 4 (57.1%) |
| Sarcomatoid | 2 (40.0%) |
| Rhabdoid | 1 (14.3%) |
| **Oncology** |  |
| Clinical T substage at Dx |  |
| 1 | 2 (20.0%) |
| 2 | 2 (20.0%) |
| 3 | 3 (30.0%) |
| 4 | 3 (30.0%) |
| Clinical N substage at Dx |  |
| cN1 | 4 (40.0%) |
| Metastasis at Dx (cM1) | 6 (60%) |
| IMDC risk group |  |
| Favorable | 0 |
| Intermediate | 4 (66.7%) |
| Poor | 2 (33.3%) |
| Type of ICI |  |
| ICI+ICI | 4 (40.0%) |
| ICI+TKI | 6 (60.0%) |
| Start of ICI to Nx, months | 8.7 (3.6-11.5) |
| Continued ICI post-Nx | 4 (57%) |
| Months on ICI post-Nx (n=7) | 1.3 (0.0-7.5) |
| **Radiology Assessment** |  |
| Size prior to ICI (cm) (n=8) | 9.8 (6.1-14.7) |
| Size prior to Nx (cm) (n=9) | 6.4 (5.5-10.3) |
| Change in size (cm) (n=7) | -4.8 (-5.2 – -1.7) |
| Change in size (%) (n=7) | -43.7 (-47.6 – -18.7) |
| **Pathology Assessment** |  |
| Laterality |  |
| Left | 1 (10.0%) |
| Right | 9 (90.0%) |
| ccRCC histology | 9 (90.0%) |
| Largest tumor bed dimension (cm)* | 6.7 (3.5-10.0) |
| Tumor bed volume (cm<sup>3</sup>)* | 162 (32.5-450) |
| **Clinical Outcomes** |  |
| Vital status |  |
| Recurrence | 0 (0%) |
| Deceased | 1 (10%) |
| Alive without recurrence | 9 (90%) |
| Median follow-up, years (IQR) | 3.1 (1.3-3.4) |

\* Includes necrotic/regression dimensions, not just viable tumor dimensions. Largest lesion taken for multifocal tumors. ccRCC: clear cell renal cell carcinoma; Dx: diagnosis; ICI: immune checkpoint inhibitor; Nx: nephrectomy; TKI: tyrosine kinase inhibitor. Numbers are specified in parenthesis when data is missing.

With a median follow-up of 3.1 years, nine patients (90%) remain alive without recurrence and one patient (10%) has died from an unrelated cause, with no cases of recurrence observed. These findings underscore that even patients with advanced stage, intermediate/poor IMDC risk, and high-grade or sarcomatoid tumors can achieve profound and durable responses to ICI therapy, highlighting complete pathologic response as a robust surrogate endpoint for favorable long-term outcomes.

### Multivariable analysis identifies independent predictors of clinical outcome

We observed that patients who demonstrated radiologic tumor shrinkage also exhibited greater iPR%, irrespective of the baseline tumor size, thus providing complementary information (**Suppl. Figure 4A**). Radiologic shrinkage derived from paired radiologic images at both pre-ICI therapy and pre-nephrectomy time points could therefore inform extent of response prior to nephrectomy. Because the radiologic data was available for a smaller cohort, we turned to pathology and DL-derived features that were available for most of the patients. For the DL-derived immune infiltrate we used a practical cutoff of 25% based on the DL medians of the two cohorts (one immune cell in 4 tumor cells), though similar trends were seen with median cutoff. Multivariable Cox regression showed three variables to be independently predictive of FFNT: (1) iPR% at nephrectomy, which was associated with longer FFNT (HR 0.97; 95% CI 0.95–0.99; p=0.0023); (2) lower percentage (≤25%) of immune infiltrate (HR 4.08; 95% CI 1.63–10.2; p=0.0026); and (3) tumor largest dimension at nephrectomy (HR 1.20; 95% CI 1.08–1.33; p=0.0006) (**Table 4**). Together, these results demonstrate that pre-nephrectomy radiologic assessment and pathologic measures go hand in hand and pathologic response, particularly iPR and immune infiltrate, provide independent prognostic information.

**Table 4.** Multivariate analysis of time to starting new systemic therapy after cytoreductive nephrectomy (n=58).

|  | Univariate Analysis |  | Multivariate Analysis |  |
| --- | --- | --- | --- | --- |
|  | Hazard Ratio<br>(95% CI) | *P* | Hazard Ratio<br>(95% CI) | *P* |
| Pathologic regression (%) | 0.97 (0.95, 0.99) | 0.0017 | 0.97 (0.95, 0.99) | 0.0023 |
| Immune cells (%) |  |  |  |  |
| 0-25% | 3.69 (1.56, 8.75) | 0.0031 | 4.08 (1.63, 10.2) | 0.0026 |
| 26-100% | Reference |  | Reference |  |
| Tumor largest dimension (cm) | 1.16 (1.05, 1.27) | 0.0030 | 1.20 (1.08, 1.33) | 0.0006 |

We asked if these variables could guide clinical trial design. As shown in **Figure 3**, our integrated data of the variables from different modalities suggests that the following variables could be used to stratify patients into those with excellent response and those that are not responders: (i) ≤4cm (largest tumor dimension) at nephrectomy; (b) >80% iPR; (c) ≥30% radiologic size reduction; (d) >25% immune cell infiltrate.

**Figure 3.**
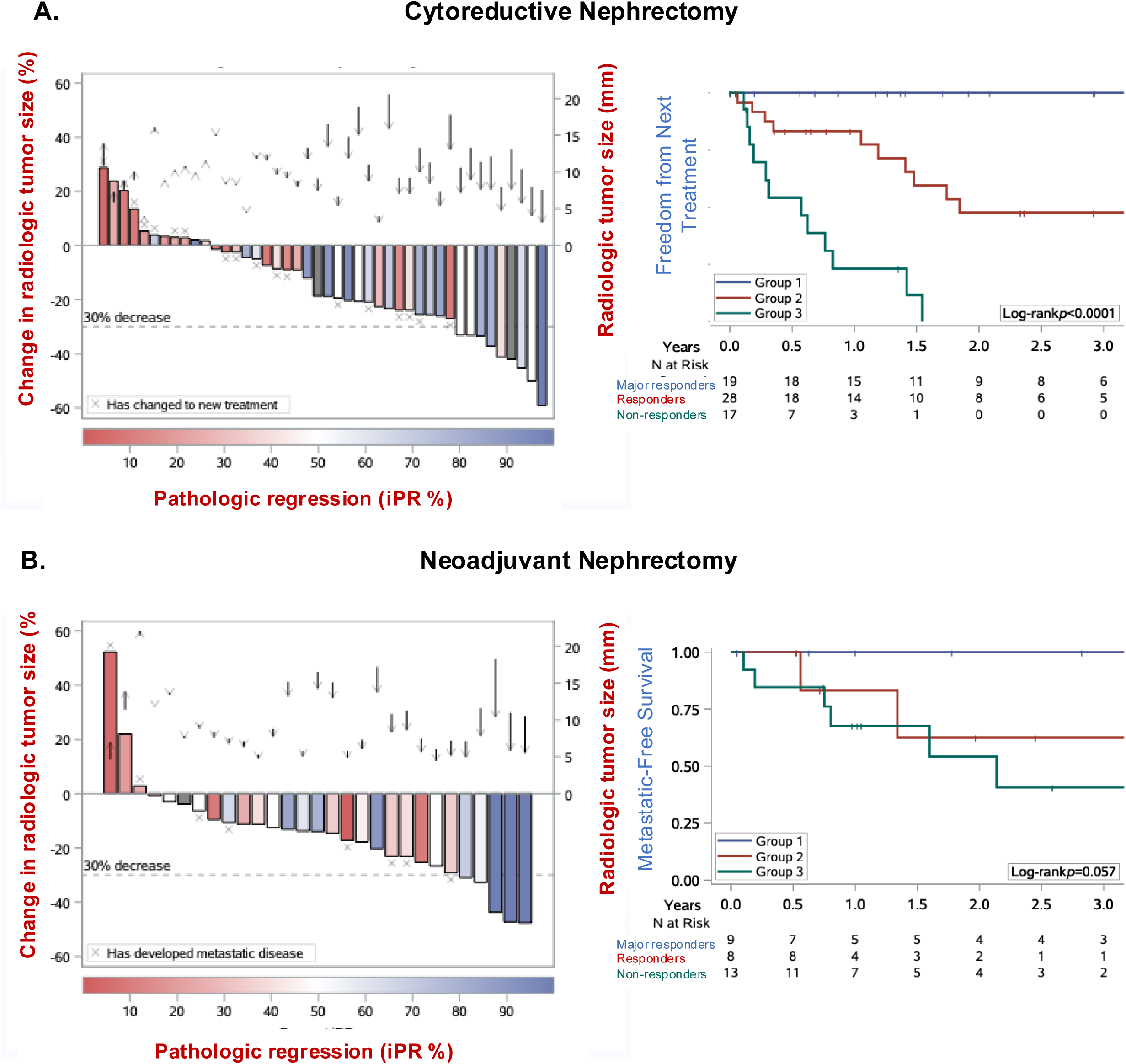
Integration of radiologic and pathologic measures of response. Waterfall plots showing patients with radiologic tumor shrinkage also demonstrated greater pathologic response, independent of baseline tumor size in both (A) CN and (B) NaN cohorts. Kaplan–Meier curves of FFNT and MSF showing stratification based on combined variables. Patients with 0-4 cm largest tumor dimension or >80% iPR had excellent prognosis across both CN and NaN cohorts (Major responders). Extent of immune cell infiltrate (responders: ≥25%; non-responders: <25%) further stratified the remaining patients in the CN cohort. Grey bars depict samples with missing iPR% (unavailable slides).

## Discussion

Unlike some other cancer types such as melanoma, breast, and lung, RCC lacks consensus criteria for pathologic response assessment in the neoadjuvant setting (9, 26, 27). In these organ systems, pathological response criteria have been shown to be superior to clinical response assessment with acceptable interobserver variability (28–30). To the best of our knowledge, this is the first work to evaluate multiple modalities and DL approaches to assess early biomarkers of response to ICI therapy in RCC. This study demonstrates that radiology, pathology, and digital pathology-based deep learning (DL) assessments provide complementary yet distinct insights into treatment response following ICI-based therapies in RCC. While radiology captures dynamic changes in tumor size, pathology provides biological depth by quantifying viable tumor and regression-associated changes, and DL enables reproducible, scalable assessments, including of immune infiltrates across large tumors. Together, this work provides a framework for integrated, standardized surrogate endpoints that can guide perioperative management and clinical trial design.

The presence of coagulative tumor necrosis in the primary tumor is a well-established adverse prognostic feature in therapy-naïve clear cell and chromophobe RCC (31–36). Necrosis is also common in papillary RCC, although it has not been shown to consistently correlate with outcome (34). The prognostic impact of the extent of coagulative necrosis has been variably reported; some studies have identified >20% necrosis as an indicator of poor survival (37), whereas others noted that extensive necrosis in low-grade RCC as being associated with a more favorable prognosis (38). In the post ICI-therapy nephrectomy setting, distinguishing therapy-induced regression (fibrosis, hyalinization, ischemic-type necrosis) from true coagulative tumor necrosis remains challenging. Because of limited awareness and the absence of standardized criteria in post ICI setting, clinical pathology reports often conflate these patterns where some include both coagulative necrosis and regression changes under the umbrella of “necrosis”, while others restrict reporting to true “coagulative necrosis” as recommended in untreated specimens. Harmonized definitions and standardized reporting will therefore be essential to reduce variability and enable multicenter comparison. Our findings demonstrate that the presence and extent of coagulative necrosis continue to confer poor prognosis even after ICI-therapy. In contrast, pathologic response correlated strongly with favorable outcomes, and these results were reproducible using unbiased DL models. Given that coagulative necrosis typically represents only a small fraction of the tumor, and that viable tumor was equally predictive, quantifying and reporting the extent of viable tumor may also serve as a practical and reproducible metric in routine practice.

This study illustrates that known prognostic features may not have the same adverse association in the post-ICI setting. Unlike the association with poor outcomes in treatment-naïve setting, presence of “necrosis” and immune infiltrate correlated with favorable outcomes when present in post-ICI nephrectomy, while high stage, presence of sarcomatoid change and high grade, historically strong predictors of poor survival, lost prognostic significance. These observations align with the paradigm shift seen in prior studies, where histologic features carry different prognostic meaning compared to untreated tumors (12). Our findings emphasize the need for a distinct framework of prognostic markers in post-ICI RCC, rather than extrapolating from untreated disease.

Our findings suggest that even partial tumor submission can adequately estimate response if sampling captures gross heterogeneity, which is practical for routine pathology workflows. Due to the large size of renal primary tumors pathological response assessment will need to be by approximation only, and submission of entire slabs may not be practical in all routine settings. This study illustrates that partial submission and just two representative slides per case evaluated by DL models may suffice, yielding results concordant with central pathology review. Further, results from these practical strategies mirrored the threshold of ≤20% viable tumor that was proposed in a recent study that evaluated entire tumor slab (12). A ≤20% viable tumor has also been widely used as surrogate endpoints in other organ types (25, 39, 40). Our results suggest that similar cutoffs may be applicable to RCC, enabling both feasibility in routine practice and comparability across studies. Importantly, DL models reproduced these findings, providing a scalable solution for multicenter implementation. Whether pathologic response within venous thrombus or metastases should be included remains unresolved; in this study, we restricted analyses to the primary tumor.

Radiology offers a temporal measure of tumor shrinkage that significantly correlated with FFNT in patients undergoing cytoreductive nephrectomy. However, radiology cannot capture the full extent of response induced by immunotherapy. Size-based assessments are often inaccurate surrogates of tumor viability: a radiographically stable tumor by size may harbor extensive necrosis, while radiologic shrinkage may underestimate viable tumor burden. The routine use of multiphasic imaging including non-contrast and contrast enhanced acquisitions on CT and MRI can assist in detecting persistent viable tumor (i.e., enhancing) and tumor induced necrosis (i.e., decreased enhancement) after treatment. Unfortunately, the variability of the acquisition protocols in our two cohorts prevented further analysis. Nevertheless, radiology also has limited capability to reliably detect pT3a and often underestimates stage, with pathologic measurements serving as the gold standard. Conversely, pathology allows detailed characterization of treatment effects, including quantification of viable tumor, fibrosis, and hyalinization (which were associated with outcome in the CN cohort), but represents a single time point and is subject to sampling variability, particularly in large or heterogeneous tumors. DL models overcome subjectivity and increase resolution by enabling reproducible quantification of tumor viability and immune infiltrate, with performance comparable to central pathology review. Thus, each modality offers distinct strengths and weaknesses, emphasizing their complementary value. Our findings underscore the importance of combining radiology for temporal dynamics, pathology for biological depth, and DL for scalable, objective quantification, thus leveraging the strengths of each discipline.

In this real-world cohort of advanced or metastatic RCC patients that received ICI before CN, >80% iPR, ≤4 cm pathologic tumor largest dimension at nephrectomy, or ≥30% radiologic size shrinkage predicted durable response, and >25% immune cell infiltrate stratifying those patients without one of those features. These findings underscore their potential as early prognostic surrogates. This is timely given the absence of standard criteria to assess pathological response in RCC and the increasing interest in adapting surrogate endpoints such as major pathological response from trials in melanoma and non-small cell lung cancer (13, 41). The integration of disease specific cutoffs in treatment decisions may inform future RCC trials designs more appropriately. However, the small neoadjuvant cohort limits definitive conclusions. Larger prospective trials are needed to validate these early biomarkers and define their role in treatment adaptation.

Although progression at metastatic sites ultimately determines outcomes, pathologic and radiologic responses in the primary tumor appear to mirror effective immune engagement against disseminated disease. Accordingly, early changes in the primary tumor may provide a practical and biologically meaningful indicator of overall therapeutic efficacy. Our findings suggest that short-term responses in the primary tumor may serve as a surrogate for long-term systemic benefit post ICI therapy. However, whether findings from the dual ICI therapy apply to patients receiving monotherapy remains uncertain.

This study has several limitations. First, its retrospective, single-institution design introduces the potential for selection bias. Second, the CN and NaN cohorts had distinct therapy choices based on treatment goal/intent and influenced timing of surgery. Third, the NaN cohort was relatively small, limiting statistical power and precluding definitive conclusions. Fourth, pathology assessment remains constrained by sampling variability and the lack of standardized response criteria. Fifth, radiologic evaluation was restricted to conventional size-based measurements, without incorporating contrast-enhanced imaging, which may better capture treatment-induced changes. Furthermore, decisions regarding systemic therapy were physician-dependent, introducing heterogeneity into treatment patterns and follow-up. Despite these limitations, the study benefits from centrally reviewed pathology, treatment at a single institution by a group of expert genitourinary cancer providers, standardization through DL models, and consistent radiology oversight.

**In summary**, our findings support the integration of radiology, pathology, and DL-based digital pathology for assessing ICI response in RCC. This study highlights the need to differentiate coagulative type tumor necrosis from pathologic tumor regression, redefine prognostic markers in the post-ICI setting, and adopt practical, reproducible measures of viable tumor burden. If validated prospectively, these approaches could inform adaptive treatment strategies and lead to earlier clinical trial endpoints in RCC.

## Data Availability

All data produced in the present work are contained in the manuscript

## Funding Sources

This work was supported by the NIH sponsored Kidney Cancer SPORE grant (P50CA196516) and endowment from Jan and Bob Pickens Distinguished Professorship in Medical Science and Brock Fund for Medical Science Chair in Pathology. These funding sources provided salary support for members of the Kidney Cancer Program (P.K.; A.C; J.M; J.B) that enabled data collection and analyses presented in this work.

### Acknowledgments

We acknowledge the patients whose samples/data provided the foundation for this study and are grateful to the Kidney Cancer Program and the Clinical Data Warehouse teams for their support and assistance.

---

Suppl. Figure 1. Representative radiologic measurements showing size shrinkage. (A) Example of baseline radiologic measurement and pre-nephrectomy primary tumor size showing tumor shrinkage.

Suppl. Figure 2. Coagulative necrosis is associated with poor prognosis in both CN and NaN cohorts. (A) Representative H&E images illustrating classic coagulative necrosis defined as homogeneous, well-demarcated zones of amorphous cellular debris with preserved outlines and karyorrhexis. (B) Pearson correlation plots showing concordance between central pathology review, DL model estimates (from 2 WSI), and standard pathology reports for coagulative necrosis. (C) Kaplan–Meier curves showing significant separation between patients with tumor that have coagulative necrosis versus those that did not, as observed during central review for FFNT and MFS. (D) Extent of coagulative necrosis significantly correlate with tumor grade and ypT substage in the post ICI therapy nephrectomy.

Suppl. Figure 3. Morphologic spectrum of ICI therapy-induced pathologic response. (A) Gross pathology specimens with mapping of viable and non-viable areas that were used to complement slide-based assessment. Blue lines depict tumor bed; red depict viable tumor; and yellow tumor necrosis. (B) Representative H&E images illustrating pathologic regression features in ICI-treated tumors (iPR). (a) Ischemic-type necrosis with hyalinized, acellular areas lacking ghost outlines, often associated with infarction. Areas of hemorrhage were not included (red line). (b-c) ICI-therapy-induced early pathologic regression including fibrin deposition, and inflammation. (d) Late therapy-related pathologic changes including fibrosis.

Suppl. Figure 4. Cross-method correlations in pathologic assessment and quantitative pathology regression estimates from deep learning (DL) and central review as correlates of treatment response. (A left) Pearson correlation plots showing concordance between central pathology review and DL model estimates for pathologic regression (iPR) demonstrating high concordance between central review and DL model estimates, despite evaluation of only two representative slides per case. (A right) Pearson correlation plots showing concordance between central review iPR% and radiologic size shrinkage. (B) Kaplan–Meier curves with stratification based on DL- based quantification of pathologic regression (iPR%) using median cutoff, showing significant differences in FFNT but not NaN.

**Supplemental Table 1.** Baseline characteristics of patients receiving cytoreductive or neoadjuvant nephrectomy after at least one dose of ICI-containing systemic therapy

|  | **Cytoreductive cohort** | **Neoadjuvant cohort** | **P** |
| --- | --- | --- | --- |
| Total | n = 66 | n = 33 |  |
| Age at Systemic Treatment | 62.5 (57-67) | 62 (58-71) | 0.4 |
| Female Sex | 16 (24%) | 8 (24%) | 0.9 |
| White Race | 55 (83%) | 28 (88%) | 0.6 |
| Hispanic Ethnicity | 10 (16%) | 6 (18%) | 0.8 |
| **Oncology** |  |  |  |
| Clinical T stage at Dx |  |  |  |
| 1 | 10 (16%) | 2 (6.1%) | 0.5 |
| 2 | 13 (21%) | 7 (21%) |  |
| 3 | 30 (49%) | 19 (58%) |  |
| 4 | 8 (13%) | 5 (15%) |  |
| IMDC risk group |  |  |  |
| Favorable | 0 (0%) | 0 (0%) | 0.9 |
| Intermediate | 28 (74%) | 17 (74%) |  |
| Poor | 10 (26%) | 6 (26%) |  |
| Type of ICI |  |  |  |
| ICI+ICI | 39 (59%) | 8 (24%) | 0.0011 |
| ICI+TKI | 27 (41%) | 25 (76%) |  |
| Start of ICI to Nx, months | 6.1 (3.7-9.5) | 4.9 (3.4-6.2) | 0.0062 |
| **Radiology** |  |  |  |
| Size prior to ICI (cm) (n = 80) | 10.4 (8.0-12.4) | 8.8 (7.1-11.8) | 0.5 |
| Size prior to Nx (cm) (n = 87) | 8.7 (7.1-11.5) | 7.6 (5.6-11.7) | 0.7 |
| Change in size (cm) (n = 73) | -1.5 (-3.5-0.0) | -1.3 (-2.3 - -0.6) | 0.9 |
| Change in size (%) (n = 73) | -18.7 (-25.8-0.2) | -13.9 (-25.3 - -9.5) | 0.9 |
| **Pathology at Nx** |  |  |  |
| Laterality |  |  |  |
| Horseshoe | 0 (0%) | 1 (3.0%) | 0.096 |
| Left | 30 (45%) | 9 (27%) |  |
| Right | 36 (55%) | 23 (70%) |  |
| Multifocal | 3 (4.5%) | 2 (6.1%) | 0.9 |
| Largest dimension, cm* | 8.1 (6.5-10.7) | 8.2 (5.5-12.4) | 0.9 |
| Volume, cm<sup>3</sup>* | 289 (158-662) | 356 (140-710) | 0.9 |
| ccRCC histology | 59 (89%) | 28 (85%) | 0.5 |
| Grade |  |  |  |
| G2 | 6 (10%) | 8 (32%) | 0.021 |
| G3 | 28 (48%) | 11 (44%) |  |
| G4 | 24 (41%) | 6 (24%) |  |
| Sarcomatoid | 13 (21%) | 4 (13%) | 0.4 |
| Rhabdoid | 11 (18%) | 5 (17%) | 0.9 |
| Coagulative Necrosis | 44 (69%) | 21 (70%) | 0.9 |
| Pathological T stage at Nx |  |  |  |
| ypT0 | 6 (9.1%) | 4 (12%) | 0.3 |
| ypT1 | 7 (11%) | 3 (9.1%) |  |
| ypT2 | 5 (7.6%) | 5 (15%) |  |
| ypT3 | 40 (61%) | 21 (64%) |  |
| ypT4 | 8 (12%) | 0 (0%) |  |
| ypN1 at Nx | 10 (15%) | 3 (9.1%) | 0.4 |
| Deceased | 17 (26%) | 5 (15%) | 0.2 |
| Median follow-up, years (IQR) | 2.5 (1.0-3.6) | 2.1 (1.2-4.1) | 0.9 |

\* Includes necrotic/regression dimensions, not just viable tumor dimensions. Largest lesion taken for multifocal tumors. ccRCC: clear cell renal cell carcinoma; Dx: diagnosis; ICI: immune checkpoint inhibitor; Nx: nephrectomy; TKI: tyrosine kinase inhibitor. Numbers are specified in parenthesis when data is missing.

