## Supplemental Figures for "Radiologic, Pathologic, and Deep Learning Predictors of Response to Immune Checkpoint Blockade in Renal Cell Carcinoma Patients Undergoing Post-Treatment Nephrectomy"

### Supplementary Figure 1

**A** **Radiologic ICI therapy related changes**

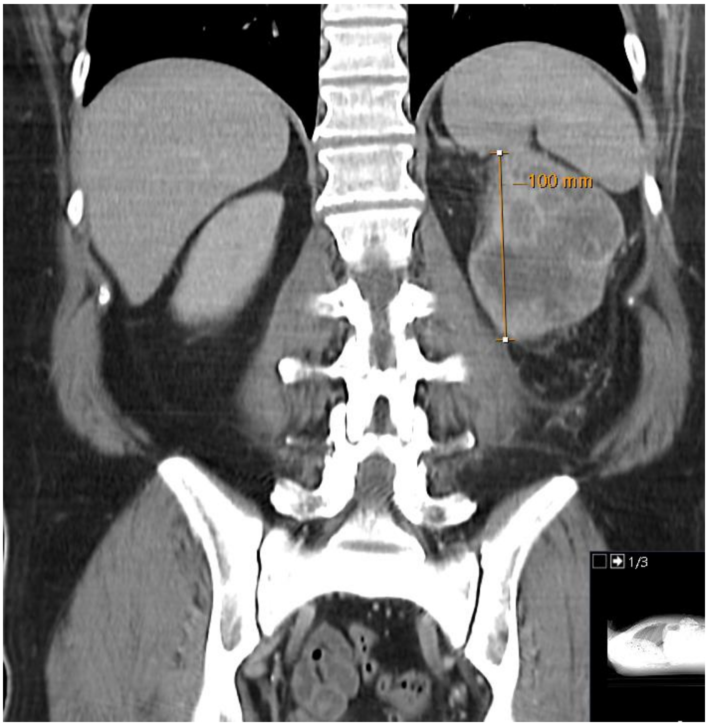


Prior to start of systemic therapy

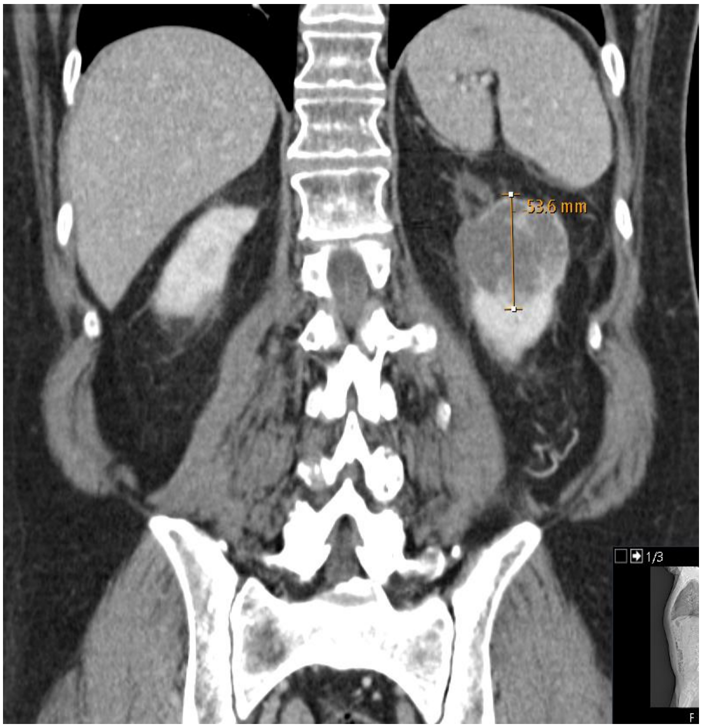


Prior to nephrectomy

**Supplementary Figure 2**

**A** **Coagulative Tumor Necrosis**

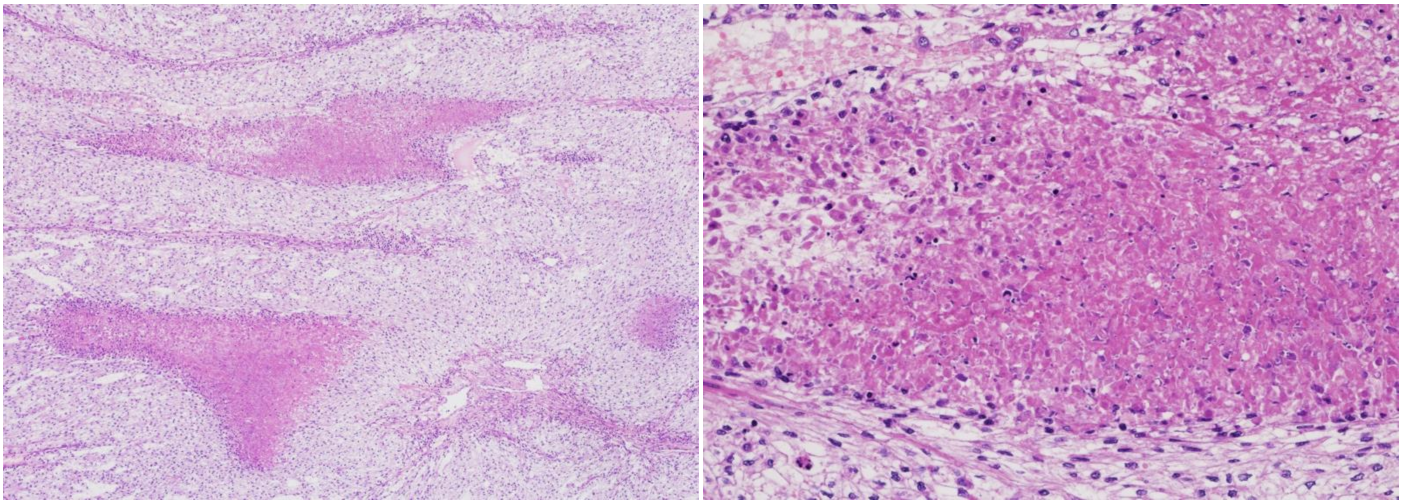

**B** **Correlation Between Central Review, Deep-Learning Model, and Pathology reports**

**Coagulative Necrosis (%)**

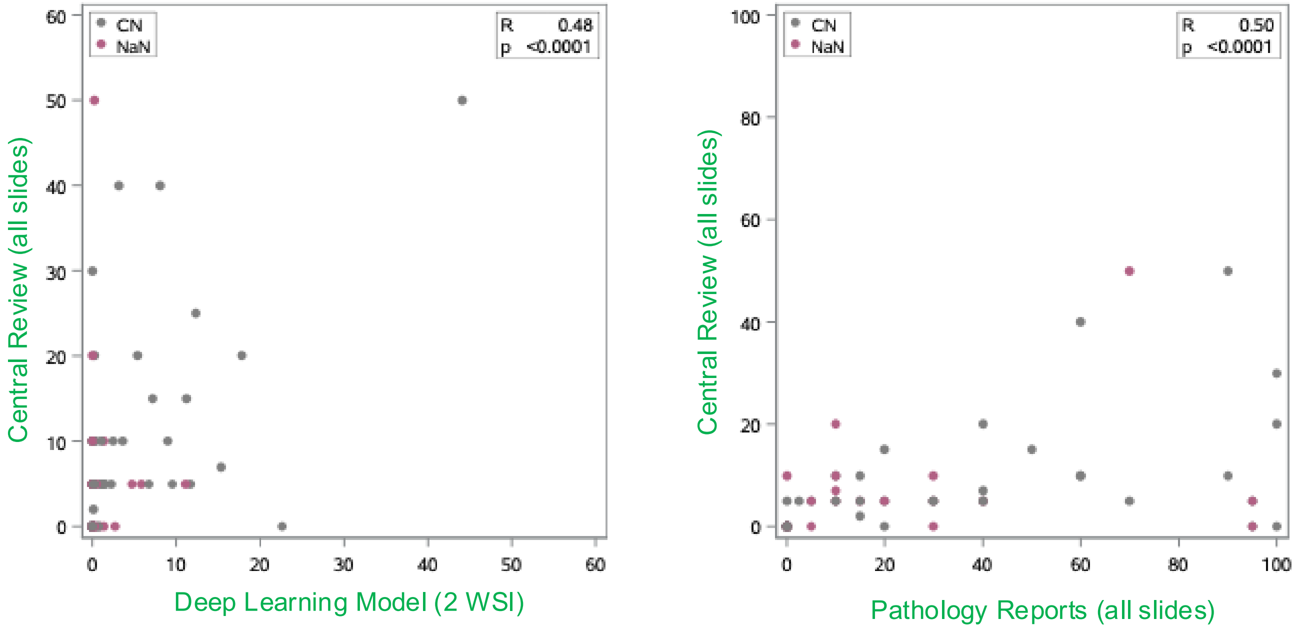

Supplementary Figure 2

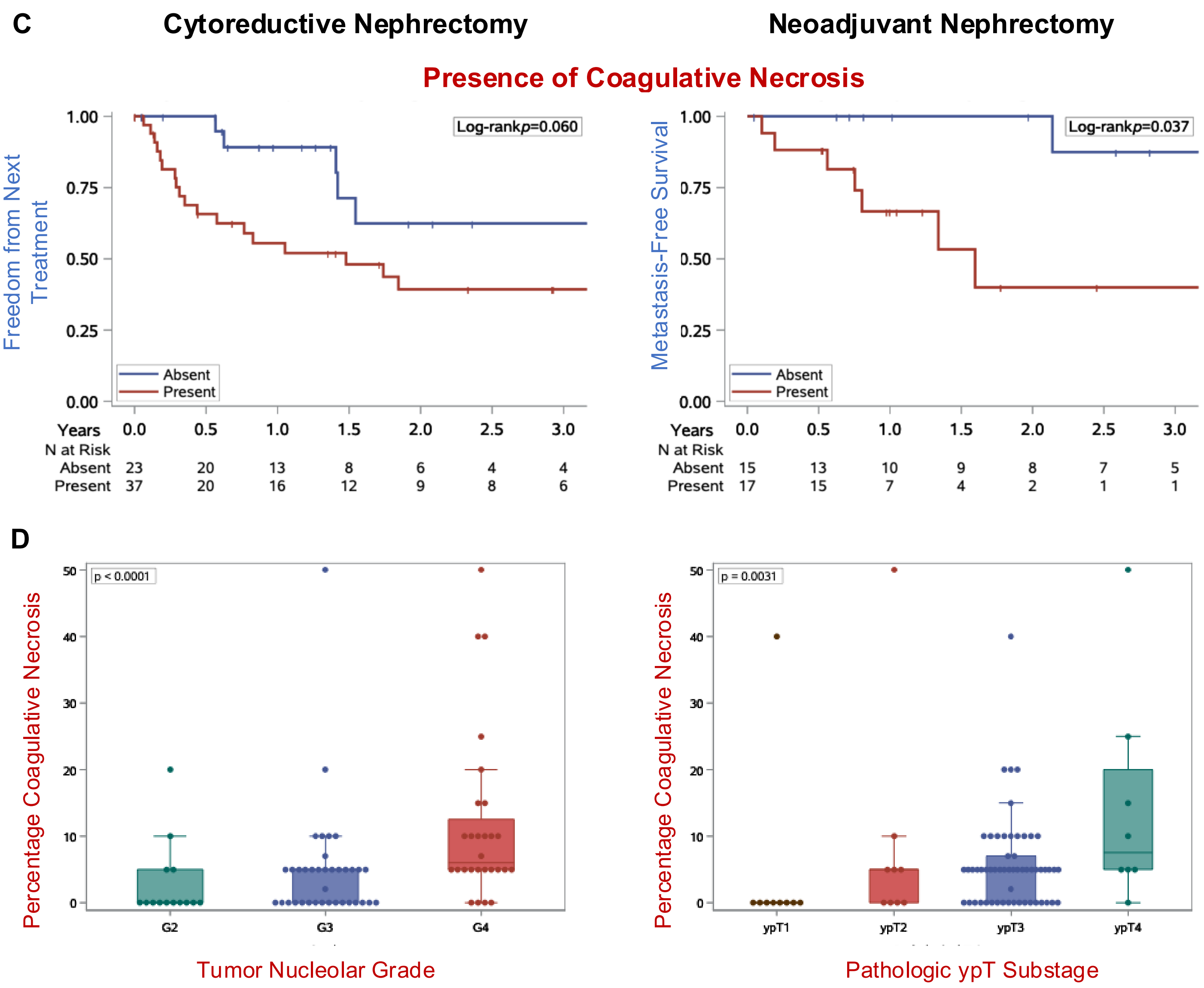

**Supplementary Figure 3**

**A** **Macroscopic ICI therapy related changes**

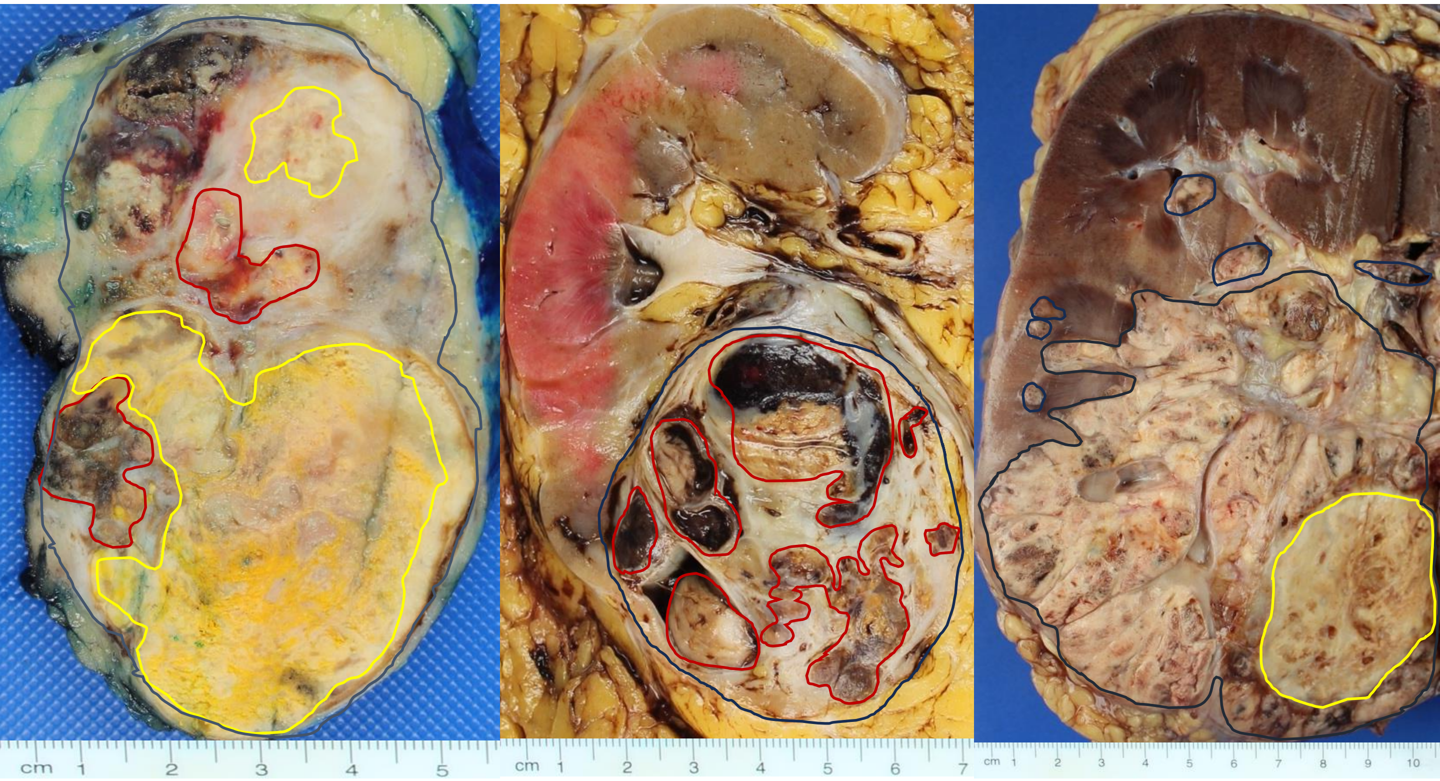


**B** **Microscopic ICI therapy related changes**

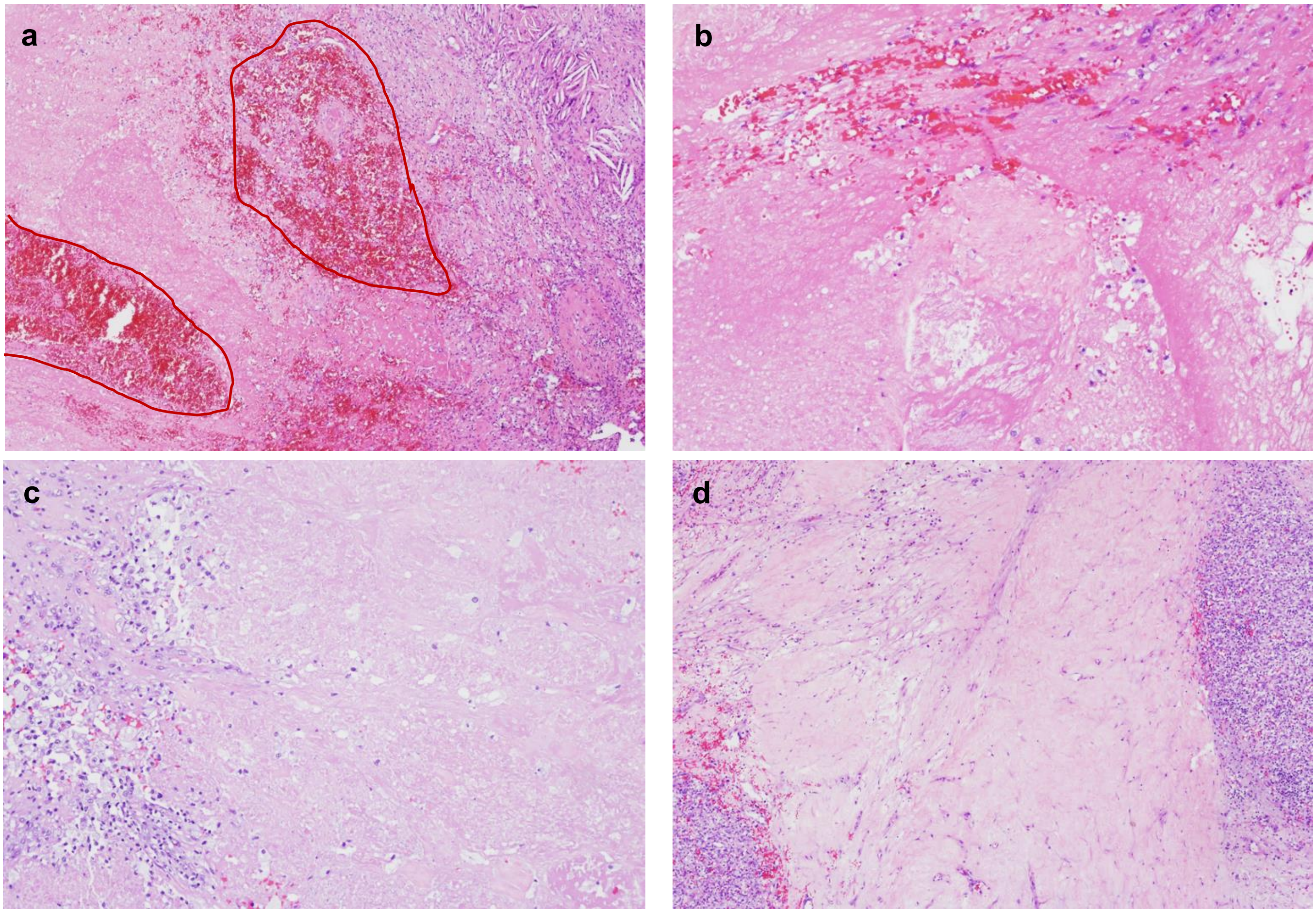

**Supplementary Figure 4**

### A. Correlation Between Pathology Reports, Central Review and Deep-Learning Model

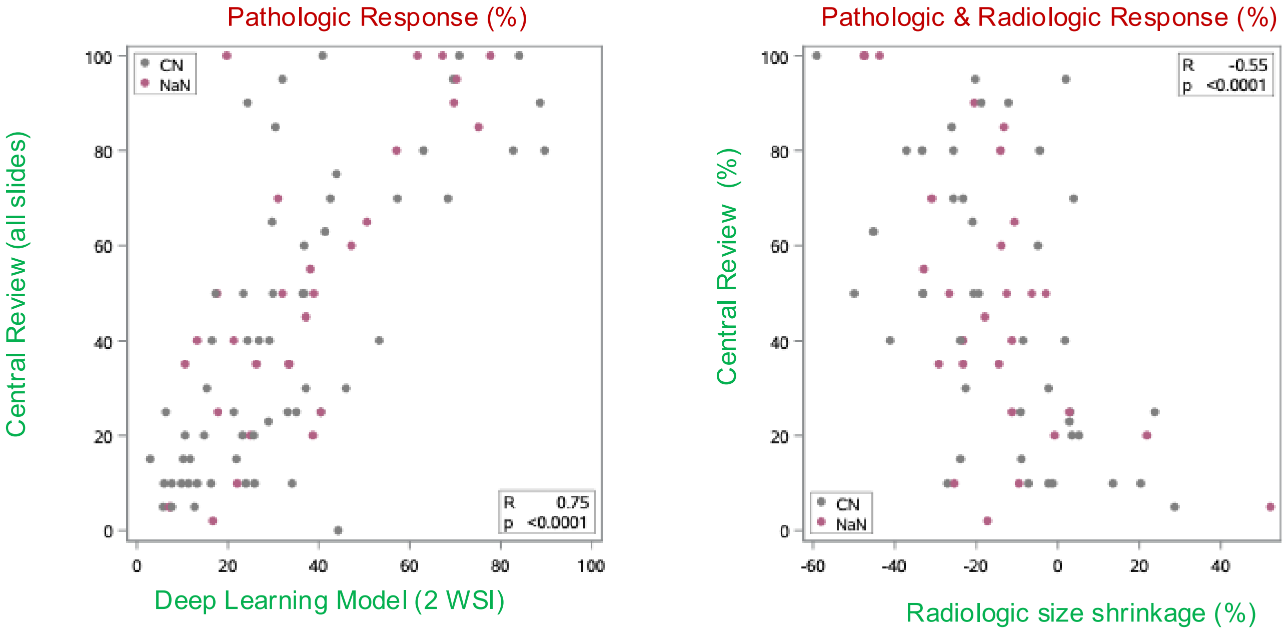


### B. Cytoreductive Nephrectomy / Neoadjuvant Nephrectomy

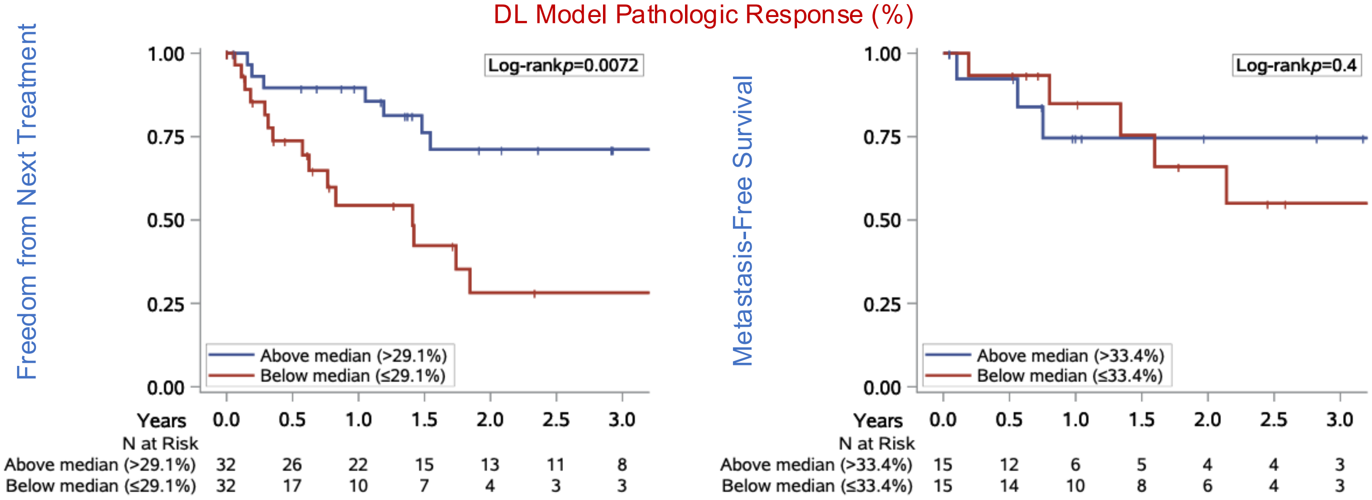
